# Incidence, risk factors and impact of seasonal influenza in pregnancy: A national cohort study

**DOI:** 10.1101/2020.07.23.20160705

**Authors:** Nicola Vousden, Kathryn Bunch, Marian Knight, the UKOSS influenza Co-Investigators Group

## Abstract

**Background:** Pregnant women are particularly vulnerable to severe infection from influenza resulting in poor neonatal outcomes. The majority of evidence relates to pandemic 2009 A/H1N1 influenza. The objective of this study was to describe the characteristics and outcomes of pregnant women hospitalised with seasonal influenza.

**Methods:** This national, prospective, observational cohort study used the UK Obstetric Surveillance System (UKOSS) to identify all pregnant women admitted to hospital between 01/11/2016 and 31/10/2018 with laboratory confirmed influenza together with a comparison group of pregnant women. Baseline characteristics, immunisation status, maternal and perinatal outcomes were compared.

**Results:** There were 405 women admitted to hospital with laboratory confirmed influenza in pregnancy: 2.7 per 10,000 maternities. Compared to 694 comparison women, women with influenza were less likely to be professionally employed (aOR 0.59, 95%CI 0.39-0.89) or immunised in the relevant season (aOR 0·59, 0·39-0·89) and more likely to have asthma (aOR 2.42, 1.30-4.49) or have had a previous pregnancy complication (aOR 2·47, 1·33-4·61). They were more likely to be admitted to intensive care (aOR 21.3, 2.78-163.1) and to have a caesarean birth (aOR 1·42, 1·02-1.98). Their babies were more likely to be admitted to neonatal intensive care (aOR 1.86, 1·01-3·42).

**Conclusions:** Immunisation reduces the risk of hospitalisation with influenza in pregnancy which is associated with increased risk of morbidity for both the mother and baby. There is a continued need to increase awareness of safety and effectiveness of immunisation in pregnancy and provision within antenatal care settings, especially for high risk groups.

**Key points:** The incidence of hospital admission with seasonal influenza in pregnancy in the UK is 2.7 per 10,000 maternities. Lack of immunisation increases risk of hospitalisation. Influenza in pregnancy increases risk of caesarean birth and neonatal admission to intensive care.

## Introduction

In 2012, the World Health Organization recommended that countries initiating or expanding seasonal influenza immunisation programmes should prioritise pregnant women over other high-risk groups.[1] In the UK, national guidance since 2010-11 has been that all pregnant women, at any gestation, should receive immunisation, with an ambition to reach at least 75% of this population.[2] This policy is based on increased risk of complicated influenza in pregnant women, protection of both mother and infant and the safety and effectiveness of the vaccine,[1] yet uptake varies globally.[3]

Confirmed seasonal influenza is reported to affect between 483 to 1097 pregnant women per 10,000 and between 3 to 91 infants per 10,000.[4] Evidence, predominantly from the pandemic influenza period in 2009 (H1N1), has demonstrated that pregnant women were particularly vulnerable to severe infection,[5-9] which results in increased risk of hospital admission.[10] Influenza in pregnancy has not been shown to increase the likelihood of maternal mortality or severe morbidity compared to the general population of women of reproductive age but the quality of evidence to date is very low.[10] Factors associated with increased risk of hospital admission after pandemic influenza infection in pregnancy include maternal obesity, asthma, multiparity, multiple pregnancy, black or other minority group ethnicity and smoking among women younger than 25 years.[5, 7, 9, 11] Pandemic influenza has also been widely reported to increase the risk of poor infant outcome such as preterm birth and stillbirth,[10] although more recent studies suggest this is only observed in women with underlying medical comorbidity[12] and is not observed in cases of mild influenza.[13]

Recent evidence suggests that the increased risk of hospitalisation may also be observed in pregnant women with seasonal influenza,[14, 15] which again is more likely in women of non-European ethnicity.[14] However, the lack of high-quality studies reporting seasonal influenza makes it challenging to assess the risk of associated morbidity, the characteristics of those at risk or to compare this risk to pandemic influenza and inform associated policy. Recent systematic reviews have concluded that further evidence regarding the impact of seasonal influenza on maternal and neonatal outcomes is required.[10, 16]

Influenza immunisation during pregnancy is widely reported to have no negative impact on pregnancy outcomes. Benefits in terms of reduction in risk of certain adverse pregnancy outcomes, including preterm birth,[17, 18] low birth weight[18, 19] and stillbirth[20], have been reported. Immunisation is effective at preventing seasonal influenza in pregnant women and their infants in both high and low-resource settings[21-24] and is cost-effective for both seasonal and pandemic influenza.[25-27] Despite this, uptake in pregnant women is low, with 45% of all pregnant women in England receiving immunisation in 2018-19, a reduction from 47% the year before [28]. Monitoring vaccine uptake in pregnant women is complicated by the dynamic nature of the group, coding for pregnancy status and differing place of vaccine administration, with immunisation available in primary care and some community pharmacies as well as antenatal clinics.[28] This may contribute to an underestimation of reach, but it is likely this still falls considerably short of the overall national target of 75% coverage of risk groups in the UK.

The most commonly cited reasons for not being immunised include perceptions of risk of influenza and concern over effectiveness or safety of the immunisation.[29-31] Characteristics of women associated with an increased likelihood of receiving seasonal influenza immunisation are White British ethnicity,[31] professional employment and education,[29, 32] immunisation in a previous pregnancy and receiving information around immunisation from a healthcare professional.[32] Similar characteristics were associated with uptake of pandemic flu vaccine in 2009.[33, 34]

There is thus clear evidence that pregnant women are at greater risk of influenza and that pandemic influenza is associated with negative impact on pregnancy outcomes. Seasonal influenza immunisation during pregnancy protects against influenza in women and their children and is safe and cost-effective. However, there is insufficient evidence to describe the impact of seasonal influenza on pregnancy outcomes, the characteristics of those at greatest risk and therefore the associated influence of prior immunisation. These data will inform policy and investment at a national and international level.

## Objective

The objectives of this study were to:

- To use a national maternity research platform to determine the incidence of hospitalisation with seasonal influenza in pregnancy in the UK.
- To identify the characteristics associated with hospitalisation with seasonal influenza in pregnancy.
- To determine the impact of severe seasonal influenza in pregnancy on outcomes for mother and infant.
- To identify the characteristics associated with immunisation for seasonal influenza in pregnancy.
- To determine the impact of immunisation on maternal and infant outcomes.

## Methods

A national, prospective, observational cohort study was undertaken using UK Obstetric Surveillance System (UKOSS). UKOSS is a national research platform that includes all 194 consultant-led obstetric units in the UK. Nominated clinicians are sent a monthly notification email containing a list of conditions under surveillance and are asked to report the number of women with these conditions or to confirm zero cases. In this study, reporters were asked to identify all women in the UK admitted to hospital between 1st November 2016 and 30th October 2018 with laboratory confirmed influenza infection in pregnancy. On reporting a woman with influenza, the clinician completed a data collection form with anonymised data about diagnosis and management, as well as maternal demographics, immunisation status, obstetric and medical history, delivery and perinatal outcomes. Women were excluded if a data collection form was not returned (n = 79) or they did not meet the definition of confirmed influenza (n = 41) (Figure 1). Information on women who died from influenza in pregnancy, or consequent stillbirths or neonatal deaths, was cross-checked with data from the MBRRACE-UK collaboration, the organisation responsible for maternal and perinatal death surveillance in the UK. If any additional women were identified through these sources which had not been identified through UKOSS, the nominated UKOSS clinician in the relevant hospital was contacted and asked to provide further information on management and outcomes.

**Figure 1.**
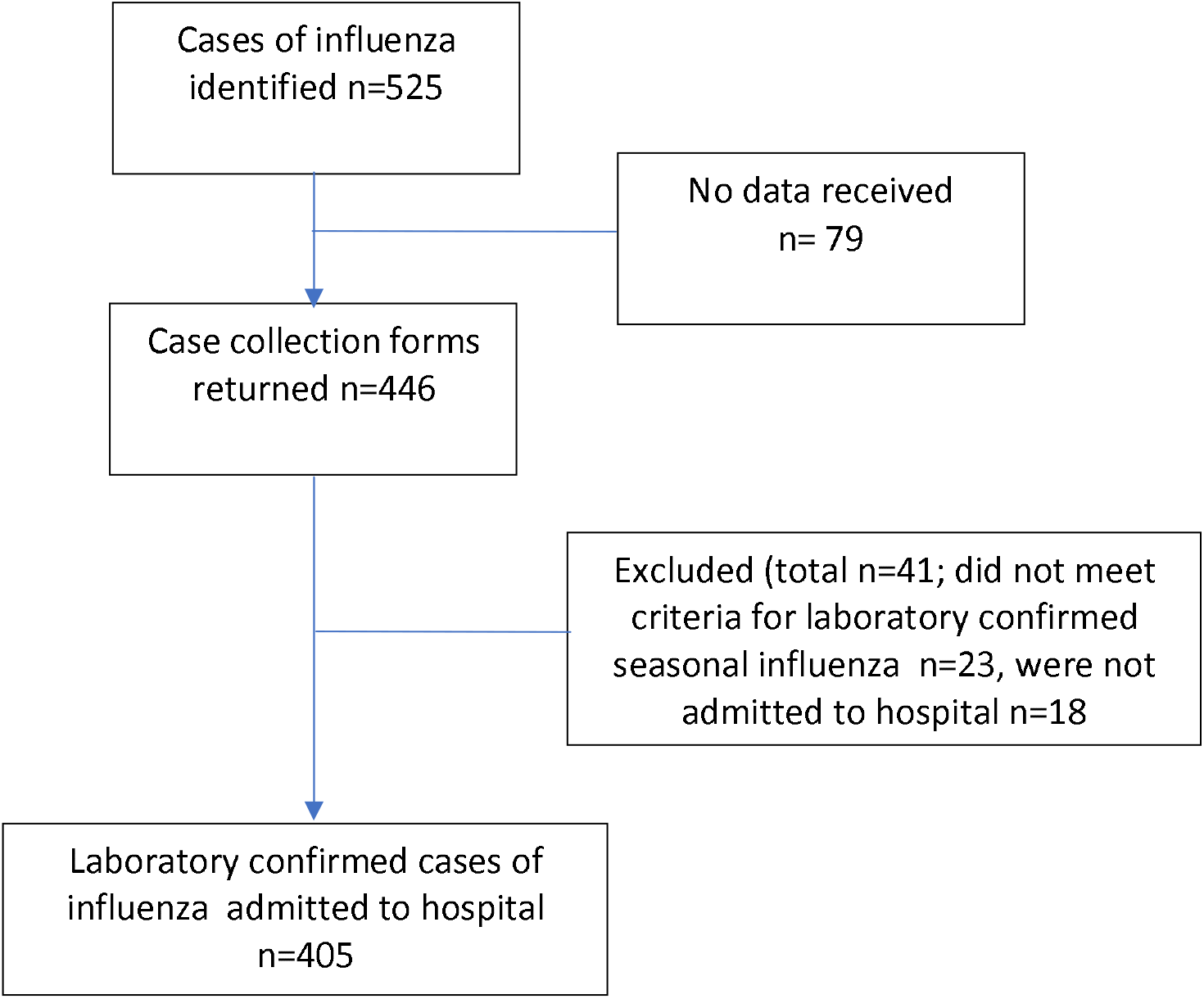
Identification of case women eligible for inclusion in the study.

Since all consultant led obstetric units report to UKOSS, this system captures all women hospitalised with influenza from the entire pregnancy population. The incidence of seasonal influenza was thus calculated using denominators from national maternity data.

The comparison group consisted of the two women that delivered in the same hospital immediately before each woman diagnosed with influenza between 1^st^ November 2017 and 30^th^ October 2018. Records were unavailable for 116 comparison women.

### Ethics committee approval

This study was approved by the North London REC1 (REC Ref. Number: 10/H0717/20).

### Statistical Analysis

The incidence of seasonal influenza was calculated using denominators from national maternity data from 2016, 2017 and 2018 for the United Kingdom. A maternity was defined as a woman giving birth to one or more children including stillborn babies at 24 weeks’ gestation or greater. Baseline characteristics were compared between exposed and comparison women, as well as by immunisation status in the comparison group, using unconditional logistic regression. Maternal and perinatal outcomes were compared between groups and adjusted for potential confounders using multivariable unconditional logistic regression. Proportions, adjusted and unadjusted odds ratios are presented with 95% confidence intervals. Statistical analyses were undertaken using STATA version 15.

## Results

In total, there were 405 women admitted to hospital with laboratory confirmed influenza in pregnancy (exposed cohort) during the study (Figure 1). Information was received for 694 unexposed comparison women (86% of those requested). There were 1,476,405 maternities during this period, therefore the overall incidence of confirmed influenza requiring hospital admission in pregnancy was 2.7 per 10,000 maternities (95% CI 2.5-3.0).

Based on analysis of data from comparison group women, women that smoked during this pregnancy were less likely to be immunised than non-smokers (aOR 0.42, 95% CI 0.25-0.71). Women with an estimated date of delivery in July to September (n=66, 9.5%), or October to December (n=46, 6·6%) were also less likely to be immunised than those expected to deliver in January to March (aOR 0·43, 0·20-0·91; aOR 0·16, 0·04-0·74 respectively) (Table 1). No association was found between maternal age, socioeconomic status, marital status, parity, body mass index or pre-existing medical conditions and current immunisation status.

**Table 1.** Characteristics associated with immunisation in control women.

| Independent Variables | Immunisation status |  |  | OR (95% CI)<sup>a</sup> | aOR (95% CI) |
| --- | --- | --- | --- | --- | --- |
|  | Current<br>N=214 | Not<br>current<br>N=264 | Unknown<br>N=216 |  |  |
|  | number (%) | number<br>(%) | number (%) |  |  |
| **Maternal age (years)** |  |  |  |  |  |
| <20 | 6 (3) | 7 (3) | 5 (2) | 1.10 (0.36-3.36) | - |
| 20-35 | 142 (66) | 183 (69) | 152 (70) | 1.00 (-) | - |
| ≥ 35 | 66 (31) | 74 (28) | 59 (27) | 1.15 (0.77-1.71) | - |
| **Socioeconomic status**<br>**(woman or partner)** |  |  |  |  |  |
| Professional | 55 (26) | 50 (19) | 38 (18) | 1.38 (0.88-2.18) | - |
| Non-professional<br>employed | 117 (55) | 147 (56) | 130 (60) | 1.00 (-) | - |
| Unemployed | 20 (9) | 35 (13) | 21 (10) | 0.72 (0.39-1.31) | - |
| Unknown | 22 (10) | 32 (12) | 27 (13) |  | - |
| **Marital Status** |  |  |  |  |  |
| Married/Cohabiting | 185 (86) | 220 (83) | 183 (85) | 1.00 (-) | - |
| Single | 27 (13) | 41 (16) | 28 (13) | 0.78 (0.46-1.32) | - |
| Unknown | 2 (1) | 3 (1) | 5 (2) |  |  |
| **Smoking Status during**<br>**pregnancy** |  |  |  |  |  |
| Did not smoke | 184 (86) | 197 (75) | 168 (78) | 1.00 (-) | 1.00 (-) |
| Smoked | 28 (13) | 64 (24) | 43 (20) | 0.47 (0.29-0.76) | 0.42 (0.25-0.71) |
| Unknown | 2 (1) | 3 (1) | 5 (2) |  |  |
| **Parity** |  |  |  |  |  |
| Nulliparous | 92 (43) | 96 (36) | 86 (40) | 1.32 (0.91-1.91) | - |
| Multiparous | 122 (57) | 168 (64) | 130 (60) | 1.00 (-) | - |
| **Body Mass Index** |  |  |  |  |  |
| Normal or overweight<br>(<30) | 162 (76) | 193 (73) | 163 (75) | 1.00 (-) | - |
| Obese (≥30) | 48 (22) | 59 (22) | 48 (22) | 0.97 (0.63-1.50) | - |
| Unknown | 4 (2) | 12 (5) | 5 (2) |  |  |
| **Multiple Pregnancy** |  |  |  |  |  |
| Yes | 8 (4) | 3 (1) | 2 (1) | 1.00 (-) | - |
| No | 206 (96) | 261 (99) | 214 (99) | 3.38 (0.89-12.90) | - |
| **Relevant pre-existing**<br>**medical condition** |  |  |  |  |  |
| Yes | 25 (12) | 32 (12) | 20 (9) | 0.95 (0.55-1.67) | - |
| No | 189 (88) | 231 (88) | 195 (90) | 1.00 (-) | - |
| Unknown | 0 (0) | 1 (0) | 1 (0) |  |  |
| **Asthma** |  |  |  |  |  |
| Yes | 10 (5) | 14 (5) | 6 (3) | 0.87 (0.38-2.00) | - |
| No | 204 (95) | 249 (94) | 210 (97) | 1.00 (-) | - |
| Unknown | 0 (0) | 1 (0) | 0 (0) |  |  |
| **Known previous** |  |  |  |  |  |

| **pregnancy complications<sup>b</sup>** |  |  |  |  |  |
| --- | --- | --- | --- | --- | --- |
| Yes | 9 (4) | 10 (4) | 16 (7) | 1.12 (0.44-2.80) | - |
| No | 205 (96) | 254 (96) | 200 (93) | 1.00 (-) | - |
| **Estimated date of delivery** |  |  |  |  |  |
| Jan – Mar | 129 (60) | 138 (52) | 102 (47) | 1.00 (-) | 1.00 (-) |
| Apr – Jun | 69 (32) | 70 (27) | 74 (34) | 1.05 (0.70-1.59) | 1.00 (0.64 – 1.56) |
| Jul – Sep | 13 (6) | 33 (13) | 20 (9) | 0.42 (0.21-0.84) | 0.43 (0.20-0.91) |
| Oct - Dec | 3 (1) | 23 (9) | 20 (9) | 0.14 (0.04-0.48) | 0.16 (0.04-0.74) |

<sup>a</sup>Odds ratio comparing current to not currently immunised groups
<sup>b</sup>Previous pregnancy complications include mental health problems, diabetes, hypertensive disorders and pre-existing medical conditions exacerbated by pregnancy together with mid- trimester loss, stillbirth, and pre-term labour or birth

**Table 2.** Characteristics of case and control women.

| Independent Variables | Number (%) of cases (n=405) | Number (%) of controls (n=694) | OR (95% CI) | aOR (95% CI) |
| --- | --- | --- | --- | --- |
| **Maternal age (years)** |  |  |  |  |
| <20 | 14 (3) | 18 (3) | 1.30 (0.64-2.65) | - |
| 20-35 | 286 (71) | 477 (69) | 1.00 (-) | - |
| ≥ 35 | 105 (26) | 199 (29) | 0.88 (0.67-1.16) | - |
| **Socioeconomic status** |  |  |  |  |
| Professional | 61 (15) | 143 (21) | 0.72 (0.51-1.01) | 0.59 (0.39-0.89) |
| Non-professional employed | 233 (58) | 394 (57) | 1.00 (-) | 1.00 (-) |
| Unemployed | 64 (16) | 76 (11) | 1.42 (0.98-2.06) | 1.23 (0.79-1.92) |
| Unknown | 47 (12) | 81 (12) |  | - |
| **Marital Status** |  |  |  |  |
| Married/Cohabiting | 327 (81) | 588 (85) | 1.00 (-) | - |
| Single | 76 (19) | 96 (14) | 1.42 (1.02-1.98) | - |
| Unknown | 2 (0) | 10 (1) |  |  |
| **Smoking Status during pregnancy** |  |  |  |  |
| Did not smoke | 317 (78) | 549 (79) | 1.00 (-) | - |
| Smoked | 86 (21) | 135 (19) | 1.10 (0.81-1.49) | - |
| Unknown | 2 (0) | 10 (1) |  |  |
| **Parity** |  |  |  |  |
| Nulliparous | 129 (32) | 274 (39) | 0.72 (0.55-0.93) | - |
| Multiparous | 276 (68) | 420 (61) | 1.00 (-) | - |
| **Body Mass Index** |  |  |  |  |
| Normal or overweight (<30 kg/m<sup>2</sup>) | 282 (70) | 518 (75) | 1.00 (-) | - |
| Obese (≥30 kg/m<sup>2</sup>) | 116 (29) | 155 (22) | 1.37 (1.04-1.82) | - |
| Unknown | 7 (2) | 21 (3) |  |  |
| **Multiple Pregnancy** |  |  |  |  |
| Yes | 12 (3) | 13 (2) | 1.60 (0.72-3.54) | - |
| No | 393 (97) | 681 (98) | 1.00 (-) | - |
| **Pre-existing medical condition** |  |  |  |  |
| Yes | 56 (14) | 77 (11) | 1.29 (0.89-1.86) | - |
| No | 348 (86) | 615 (89) | 1.00 (-) | - |
| Unknown | 1 (0) | 2 (0) |  |  |
| **Pre-existing Asthma** |  |  |  |  |
| Yes | 41 (10) | 30 (4) | 2.50 (1.53-4.07) | 2.42 (1.30-4.49) |
| No | 363 (90) | 663 (96) | 1.00 (-) | 1.00 (-) |
| Unknown | 1 (0) | 1 (0) |  |  |
| **Known previous pregnancy complications** |  |  |  |  |
| Yes | 43 (11) | 35 (5) | 2.24 (1.41-3.56) | 2.47 (1.33-4.61) |
| No | 362 (89) | 659 (95) | 1.00 (-) | 1.00 (-) |
| **Immunisation Status** |  |  |  |  |
| Ever Immunised against influenza |  |  |  |  |
| Yes | 137 (34) | 267 (38) | 0.69 (0.53-0.92) | - |
| No | 190 (47) | 257 (37) | 1.00 (-) | - |
| Unknown | 78 (19) | 170 (25) |  |  |
| Immunised in the relevant influenza season |  |  |  |  |
| Yes | 106 (26) | 214 (31) | 0.65 (0.48-0.87) | 0.62 (0.45-0.86) |
| No | 202 (50) | 264 (38) | 1.00 (-) | 1.00 (-) |
| Unknown | 97 (24) | 216 (31) |  |  |

Women hospitalised with seasonal influenza in pregnancy were less likely to be professionally employed (15% versus 21%; aOR 0.59, 0.39-0.89) and less likely to be immunised in the relevant season than comparison women (26% versus 31%; aOR 0·62, 0·45-0·86). Women hospitalised with influenza were more likely to have asthma (10% versus 4%; aOR 2.42, 1.30-4.49) and to have had a previous pregnancy complication than those in the comparison group (11% versus 5%; aOR 2·47, 1·33-4·61). There was no difference between exposed and unexposed women in age, marital status, smoking, parity, body mass index, multiple pregnancy or pre-existing medical conditions other than asthma.

Three percent of women (n=13) were admitted with flu in the first trimester, compared to 18% (n=70) in the second and 79% (n=309) third trimester. Of the 405 women with influenza, 308 (76%) had no specific influenza type recorded. Of the 97 women with a recorded strain, 17 (18%) had A/H1N1, 73 (75%) had influenza A/H3, 6 (6%) had other influenza A strains and only 1 (1%) had a non-A flu strain. The majority of women admitted with influenza received antiviral agents (n=349, 86%) with oseltamavir (Tamiflu) being most commonly used (n=348, 99.7%). The median duration of hospital admission was 3 days (IQR 2-5days), noting that for some women this included a postnatal stay potentially unrelated to their influenza.

Maternal and perinatal outcomes are shown in Table 3. Twenty one women with influenza (5%) were admitted to intensive care (ITU) as opposed to one woman in the comparison group (aOR 21·3, 2·78-163·1), with a median admission of 3 days (IQR 2-6) compared to 5 days for the comparison woman’s ITU admission. Only one woman admitted with influenza received extra-corporeal membrane oxygenation (ECMO). Just over a quarter of women with influenza gave birth during the same admission as their influenza admission (n=114, 28%). Women with influenza were more likely to have a caesarean birth (aOR 1·42, 1·02-1·98). There was a statistically non-significant increased risk of maternal morbidity in women with influenza compared to comparison women (aOR 2·23, 0·78-6·43). There were no increased odds of pregnancy loss or assisted vaginal birth in exposed compared to unexposed women.

**Table 3.**
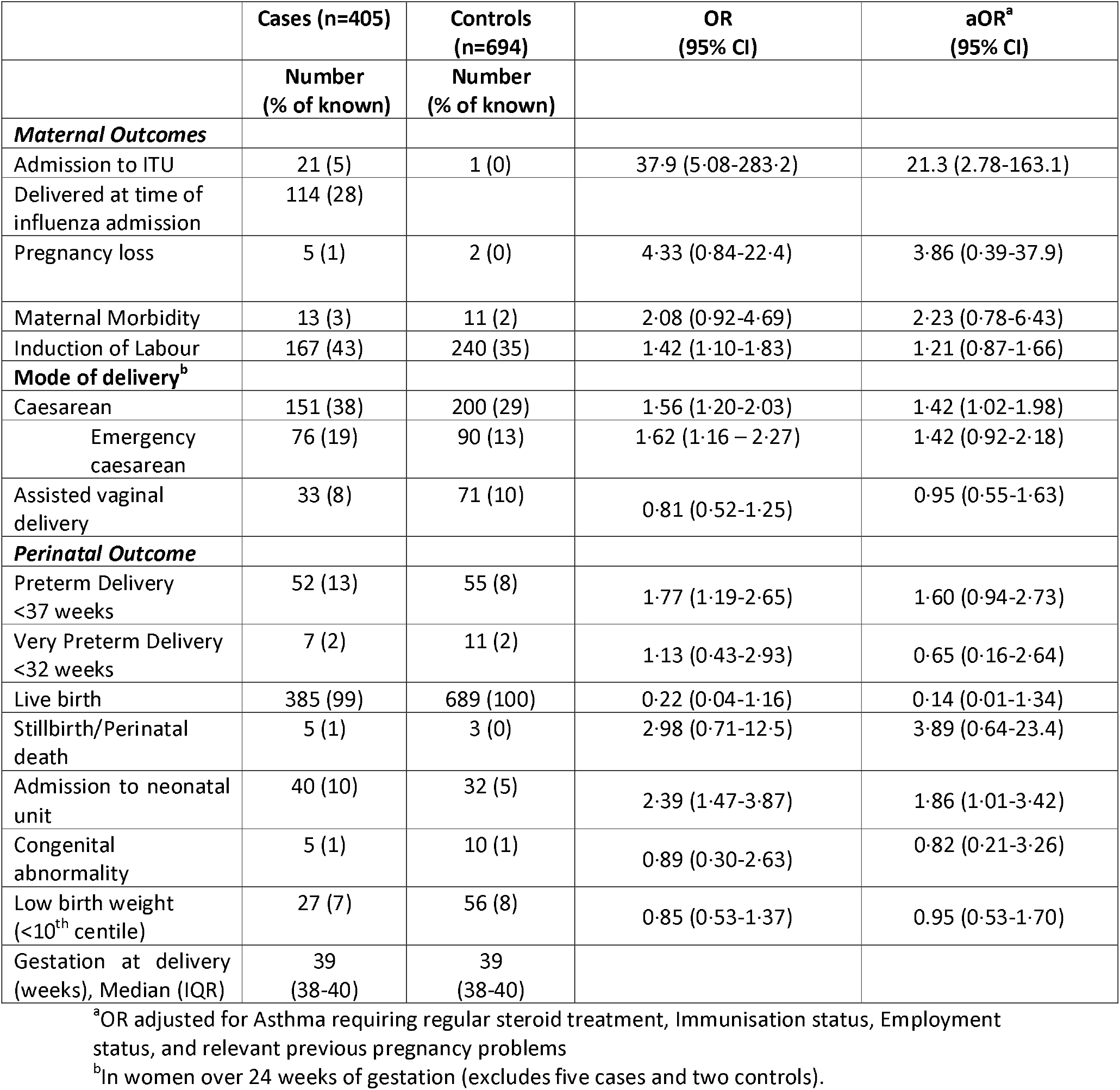
Maternal and perinatal outcomes.

The median gestation at delivery was 39 weeks both for women hospitalised with influenza and for the comparison women. A higher proportion of women with influenza gave birth at <37 weeks compared to unexposed women, but this was not statistically significant after adjustment. There was no significant difference between exposed and unexposed women in the proportion giving birth at <32 weeks. Babies of mothers with influenza were more likely to be admitted to neonatal intensive care (aOR 1·86, 1·01-3·42). Overall, there were few stillbirths, neonatal deaths and congenital abnormalities, with no differences between exposed and unexposed groups.

One hundred and six women admitted with influenza (34%) were known to have received an influenza immunisation in the relevant season. There was no difference in maternal and perinatal outcomes according to the immunisation status of the mother (Table 4).

**Table 4.** Maternal and perinatal outcomes following seasonal influenza in pregnancy outcomes by immunisation status.

|  | Immunisation status |  |  | OR (95% CI) |
| --- | --- | --- | --- | --- |
|  | Current<br>(n=106) | Not current<br>(n=202) | Unknown<br>(n=97) |  |
|  | Number (%) | Number (%) | Number (%) |  |
| **Maternal Outcomes** |  |  |  |  |
| Admission to ITU | 6 (6) | 13 (6) | 2 (2) | 0.87 (0.32-2.36) |
| Duration of ITU admission<br>(days) Median (IQR) | 2.5 (2-3) | 3 (2-7) | 4.5 (2-7) |  |
| Delivered at time of<br>influenza admission | 36 (34) | 56 (28) | 22 (23) | 1.34 (0.81-2.22) |
| Pregnancy loss | 0 (0) | 4 (2) | 1 (1) |  |
| Maternal Morbidity | 4 (4) | 7 (4) | 2 (2) | 1.06 (0.30-3.72) |
| Induction of labour | 35 (34) | 95 (49) | 37 (40) | 0.85 (0.64-1.14) |
| **Mode of delivery** |  |  |  |  |
| Caesarean overall rate | 42 (40) | 71 (37) | 38 (41) | 1.16 (0.71-1.90) |
| Emergency caesarean | 18 (17) | 40 (21) | 18 (20) | 0.80 (0.43-1.48) |
| Assisted vaginal delivery | 9 (9) | 16 (8) | 8 (9) | 1.05 (0.45-2.46) |
| **Perinatal Outcome** |  |  |  |  |
| Preterm Delivery<br><37 weeks | 16 (15) | 21 (11) | 15 (16) | 1.51 (0.75-3.03) |
| Very Preterm Delivery <32<br>weeks | 1 (1) | 3 (2) | 3 (3) | 0.62 (0.06-6.05) |
| Live birth | 103 (99) | 190 (98) | 92 (99) | 1.63 (0.17-15.8) |
| Stillbirth/ Neonatal death | 1 (1) | 3 (2) | 1 (1) | 0.61 (0.06-5.99) |
| Neonatal unit admission | 11 (11) | 19 (10) | 10 (11) | 1.08 (0.49-2.36) |
| Congenital abnormality | 2 (2) | 1 (1) | 2 (2) | 3.74 (0.34-41.8) |
| Low birth weight<br>(<10<sup>th</sup> centile) | 8 (8) | 15 (8) | 4 (4) | 0.99 (0.41-2.43) |
| Gestation at delivery<br>(weeks), Median (IQR) | 39<br>(38-40) | 39<br>(38-40) | 39<br>(38-40) |  |

## Discussion

### Main findings

We observed an incidence of hospital admission with seasonal influenza in pregnancy of 2.7/10,000 maternities, which is in keeping with hospitalisation rates reported for pandemic influenza in pregnancy. Our main findings showed that women that smoke in pregnancy and women that are due to give birth between July and December were less likely to be immunised against influenza. Women with influenza were less likely to have been immunised in the relevant season and less likely to be professionally employed than comparison women. They had a 2·5-fold increased odds of having had a previous severe pregnancy complication (e.g. hypertensive disorders of pregnancy) or asthma than comparison women without influenza. Women with influenza and their babies were more likely to be admitted to intensive care, and to have a caesarean birth compared to the unexposed group. Immunisation prior to seasonal influenza was not associated with any significant difference in maternal or perinatal outcomes.

### Strengths and Limitations

To our knowledge, this is the first national, prospective population-based study on hospital admission with seasonal influenza in pregnancy. The main strengths of this study are the robust method of nationwide prospective identification of exposed women. This allows reporting of population-based data on the causal agents and outcomes of this relatively uncommon condition. Inclusion of laboratory confirmed influenza requiring hospital admission means that only true cases were included, but we are unable to evaluate the outcomes of mild cases or those that did not present to hospital, therefore the true population incidence may be greater than reported. The comparison to a contemporaneous unexposed group allowed conclusions to be drawn on the characteristics associated with hospitalisation with influenza and immunisation and the use of camparison women from the same centres as women with influenza means there is low risk of selection or measurement bias.

A further limitation is that the time period in which this study was undertaken included a period where the influenza vaccine was less effective than previous years.[28] This may have influenced the incidence of influenza in immunised individuals as well as the risk of maternal and perinatal morbidity in this group. Some data were incomplete, the most frequent missing data was immunisation status (missing for 24% of cases and 31% of controls), as it is not routinely captured electronically in all hospitals.

### Interpretation

Prior to this study there was an absence of high-quality studies reporting maternal and perinatal outcomes following seasonal influenza.[10, 16] This study shows that women hospitalised with influenza and their babies are more likely to be admitted to intensive care, and to have a caesarean birth. We also showed that immunisation is effective at preventing influenza in pregnancy, however the overall uptake in this cohort was low, even compared to the national average of 45% in 2018-19. Increasing uptake of immunisation in pregnancy may therefore reduce this avoidable morbidity. This is particularly important in the context of services pressurised by SARS-CoV-2 infection.

The proportion of pregnant women that are reported to refuse or decline immunisation in the UK is stable at 5·7%.[28] This relatively low percentage of women who decline suggests that many women are not offered or advised to receive immunisation.[29] Indeed, it is known that there is a strong link between provision of information on immunisation by maternity healthcare workers and vaccine uptake in women.[35-37] A survey of 3441 practice nurses, midwives and health visitors working in England found that, whilst the majority stated they routinely recommend influenza immunisation in pregnancy (73%), fewer were immunised themselves (58% of midwives, 61% of health visitors and 79% of practice nurses), most commonly citing concern over side effects. Whilst midwives were identified as having the main responsibility for advising on immunisation, only 62% had received training, 60% were confident in giving advice and 9% reported providing immunisation to pregnant women, even though the majority would be happy to do so.[38] There may be a role for steps to increase healthcare professionals’ awareness about the effectiveness and safety of immunisation in pregnancy and increase provision within antenatal care settings to further prevent avoidable morbidity.

Although this study did not show increased risk of influenza in women that smoke, this group were less likely to be immunised and therefore were at greater risk of developing influenza. It is plausible that influenza may be associated with more severe respiratory complications in this group, who as a result should be a specific focus of immunisation programmes.

Women due to give birth between October and December likely face a greater risk of developing influenza in pregnancy yet they are less likely to be immunised. The main immunisation programme in the UK usually commences in November, when this group are in their third trimester. Even in their third trimester, women should be encouraged to receive immunisation at the earliest opportunity. Women due to deliver between July and September were also less likely to be immunised. This group would be in their first trimester during the influenza period, therefore this may represent the delay in recognising or booking pregnancy with a health care professional or an intentional delay through concern over the safety of immunisation in the first trimester. Women should be reassured that immunisation against influenza is safe even in early pregnancy[18] and immunisation should be encouraged at any point during the seasonal influenza period.

Women with specific previous pregnancy complications such as hypertensive disorders were more likely to get influenza but not more likely to be immunised. Hence this study highlights three high-risk groups for whom there should be a greater focus on facilitating immunisation.

Previous studies have demonstrated a reduced risk of neonatal complications following influenza in women that were immunised[17-20], whereas we did not show any significant difference in maternal or perinatal outcomes. It is possible that our study is underpowered to show a meaningful difference in this subgroup analysis. Further research is required to explore this finding.

### Conclusion

Hospitalisation with seasonal influenza in pregnancy is not uncommon, and is associated with increased risk of morbidity for both the mother and baby. We showed that influenza immunisation reduces the risk of hospitalisation with influenza in pregnancy. However, although immunisation against influenza is known to be safe, uptake of influenza immunisation in the UK remains low. There is a need to increase awareness of safety and effectiveness of immunisation in pregnancy and provision within antenatal care settings, especially for high risk groups.

## UKOSS Influenza Co-investigators Group

Peter Brocklehurst, Birmingham Clinical Trials Unit, University of Birmingham

Marian Knight, National Perinatal Epidemiology Unit, University of Oxford

Jennifer J Kurinczuk, National Perinatal Epidemiology Unit, University of Oxford

Pat O’Brien, Department of Obstetrics and Gynaecology, University College Hospitals NHS

Foundation Trust, London

Maria Quigley, National Perinatal Epidemiology Unit, University of Oxford

## Data Availability

Data may be made available following written request to the corresponding author.

## Acknowledgements

The authors would like to acknowledge the assistance of UKOSS reporting clinicians, the UKOSS Steering Committee and The NIHR Clinical Research Networks without whose support this research would not have been possible. This paper presents independent research jointly funded by the National Institute for Health Research (NIHR) Health Technology Assessment Programme (Grant reference 11/46/12) and the National Institute for Health Research (NIHR) Policy Research Programme in the Department of Health and Social Care, England (Grant reference 108/0001).

